# Identification of (ultra-)rare functional promoter mutations in cancer using sequence-based deep learning models

**DOI:** 10.1101/2025.05.06.25327057

**Authors:** Tijs van Lieshout, Carlos G. Urzúa-Traslaviña, Lucía Barbadilla-Martínez, Minh Chau Luong Boi, Harm-Jan Westra, Noud H.M. Klaassen, Vinícius H. Franceschini-Santos, Miguel Parra-Martínez, Jeroen de Ridder, Bas van Steensel, Emile Voest, Lude H. Franke

**Author notes:** Contributed equally (Bas van Steensel, Emile Voest).

## Abstract

The identification of non-coding somatic cancer-driver mutations remains challenging due to difficulties in interpreting rare and ultra-rare variants. We hypothesized that sequence-based models can be used to systematically prioritize such mutations for their functional relevance. Here we present a computational framework that leverages sequence-based models to assess the functional impact of (ultra-)rare somatic single nucleotide variants (SNVs) in promoter regions. We analysed SNVs derived from 24,529 whole-tumour genomes from three cohorts and applied the sequence-based model PARM, which was trained on massively parallel reporter assay data. We identified up to 492 promoter regions significantly enriched for putatively functional SNVs, including known cancer-drivers such as *TERT*. Overall, we find that functional promoter mutations are significantly enriched in established cancer-driver genes (*p*-value = 9.7×10^-5^). Cross-cohort validation and replication using an independent sequence-based model (Borzoi) identified nine candidate cancer genes where the prioritized promoter mutations were shown to be functional by affecting gene expression levels. These genes included well known cancer genes such as including *TERT, TP53* and *PMS2*, but also several new candidates for which no coding mutations have previously been implicated in cancer, including *PMS2, AIMP2, SASS6, RPL13A, ALKBH4, FICD* and *YAE1*. These findings demonstrate the utility of sequence-based models for identifying functional non-coding mutations and provide a framework for uncovering regulatory elements implicated in cancer.

## Introduction

The study of tumour genomes has historically focused on protein-coding regions, which comprise 3% of the genome. Recurrently mutated genes, mutated hotspot regions, and mutated single base pair changes that drive cancer have been identified in coding regions of tumour genomes (1). However, cancer risk GWAS loci have also revealed many non-coding variants (2), highlighting the importance of also studying the non-coding genome in cancer as well. With the notable exception of the *TERT* promoter, the Pan-cancer Analysis of Whole Genomes (PCAWG) consortium found that recurrent base pair changes in non-coding regions were much rarer than their coding counterparts (3). Yet, statistical enrichment analysis of non-coding mutations using sliding genomic regions of 1−10 kb pointed to potential new regulatory elements that act as candidate cancer drivers (4). However, a major issue that limits the power of these statistical approaches is that most somatic variants in cancer are passenger variants (i.e. those variants that likely have no consequence) that are not being positively selected which decreases the signal-to-noise ratio. A further challenge with these statistical approaches is the highly variable background mutation rate across cancer genomes, which requires careful corrections, and has led to exclusion of tumours with hypermutated genomes like melanoma and lymphoma to be excluded from analysis into non-coding regions (4).

We reasoned that statistical power could be gained by focusing on non-coding mutations that are likely to impact gene mRNA expression, further increasing the signal-to-noise ratio. This strategy could be further enhanced by taking the direction of the regulatory effect (up- or down- regulation of a nearby gene) into account, as this would allow the identification of clusters of neighbouring non-coding mutations with consistent effect directions. This approach would require prior knowledge of the regulatory effects of non-coding mutations. It is however difficult to determine these regulatory effects using for example RNA-sequencing, due to the low recurrence of non-coding somatic mutations.

Recently, several sequence-based models were reported to reliably predict, among other outcomes, the mRNA expression consequences of non-coding single-nucleotide variants (SNV), especially near the transcription start site (5–7). For example, we developed *Promoter Activity Regulatory Model* (*PARM*), a sequence-based model trained on massively parallel reporter assay data to predict autonomous promoter activity at genome-scale (8). An alternative model, *Borzoi*, has a bigger genomic window and was trained on RNA-seq and epigenome data (9). We thus hypothesized that the predictions of these models could help to determine if specific promoter regions in cancer genomes are enriched for putatively functional somatic mutations. In contrast to previous approaches, this approach could also be leveraged to study hypermutated tumours, since passenger variants and functional variants could be distinguished by the model predictions, irrespective of the mutation load.

In this study, we collected somatic SNV calls within promoter regions of tumour genomes of patients with cancer from the Genomics England 100,000 Genomes Project (GEL; *n*=11,073 whole-genome sequencing (WGS) samples), Hartwig Medical Foundation (HMF; *n*=7,171 WGS samples) and the International Cancer Genome Consortium (ICGC; *n*=6,285 WGS samples) (10–12). We first tested the GEL dataset for enrichment of putatively functional variants in promoter regions using PARM, resulting in the identification of 247 candidate driver promoter regions. We subsequently reproduced these hits using *Borzoi* and through replication in the independent HMF and ICGC cohorts, which resulted in 50 reproducible candidate driver promoter regions. Using paired RNA-seq data of cancer patients in the HMF cohort, we established for nine of these regions that the prioritized variants significantly affect gene expression levels. Finally, we identify many additional putative non-coding cancer driver regions by meta-analysing the GEL, HMF and ICGC datasets. These results show how sequence-based model predictions can be leveraged to accelerate the identification of non-coding regions and their possible variants that could act as cancer drivers. In line with this goal, we provide the resulting summary statistics as a resource with this manuscript (Table S1).

## Results

To study the functional impact of non-coding somatic mutations, we performed a mutational burden analysis in which we compared the predicted function of all the observed variants across all patients in a single promoter region against the prediction of comparable variants that are not observed in any of the other patients of that cancer cohort in the same region (**Figure 1**). We restricted our non-coding analyses to the promoter region of genes because the transcriptional consequences of distal variants remain harder to predict with currently available sequence-based models (6).

**Figure 1:**
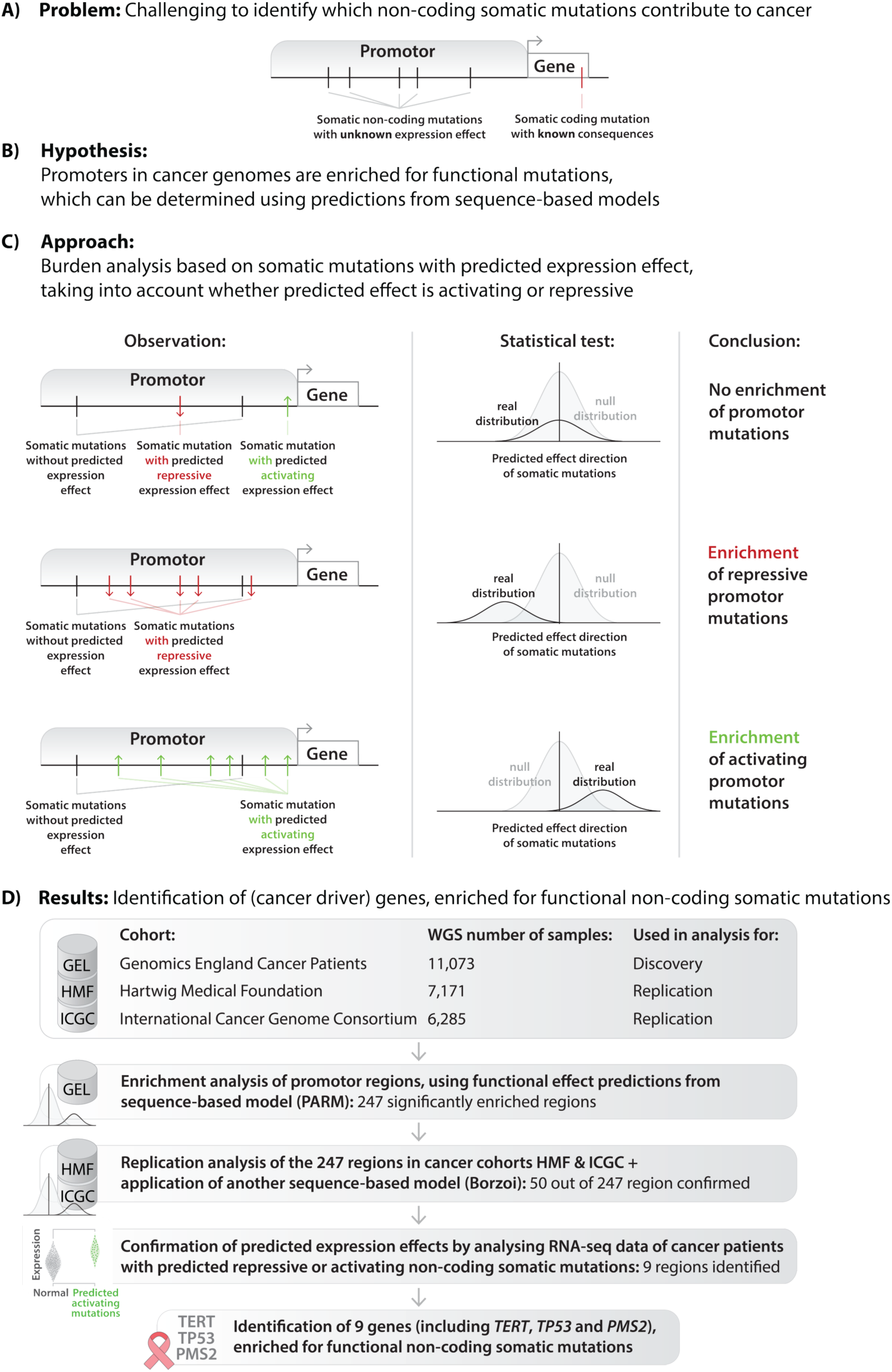
Overview of the interrogation of promoter regions to identify enrichments in functional non-coding somatic mutations.

### Burden analysis strategy using predicted function re-identifies known cancer driver genes

To determine if the enrichment of functional variants is different from chance, we compare the functional score predictions of the observed mutations against the functional score predictions of nearby but non-observed mutations. We use two null models for this: the “Alternative Alleles” and “Same Promoter” models (see Methods, Fig. S1) to capture different potential modes of action of non-coding somatic mutations. Briefly, the Alternative Alleles model assumes that functional SNVs would not be functional if an alternative allele would have occurred at the same base pair location as is the case with gain-of-function variants. For example, in the *TERT* promoter the -146 C>T somatic variant creates an ETS motif, while the C>A or C>G variants at the same position do not. Therefore, for variants in a candidate region, this null model consists of all the mutations at the same base pairs that were not observed. The Same Promoter model assumes that functional SNVs would not be functional if the same variant would have occurred elsewhere in the same promoter. For example, a mutation in a motif can often disrupt its function regardless of the substitute nucleotide but the same variant elsewhere in the promoter would not show function. Therefore, for variants in a candidate region, the Same Promoter consists of all unobserved variants in the same promoter that share the trinucleotide context with observed variants to ensure the variants are optimally comparable. As results, we considered regions that were found in both null models to maximize the power to detect both types of driver variant mechanisms (Fig. S1).

As a sanity check, we determined whether our approach would be able to re-identify known coding somatic driver genes. For this check, we conducted a somatic variant burden test with both background models, but used variants within coding regions instead of promoters and using precalculated CADD1.7 (13) scores as functional scores (derived from the Meta ESM-1v protein model, **Table 1**). This resulted in 942 genes that passed Bonferroni-significance with either of the background models (*p* < 0.05/17,168 genes = 2.91×10^-6^), including 118 genes that are already known pan-cancer and/or cancer-specific drivers (including *TP53*, *PIK3CA, BRAF, PTEN, KRAS*, *ARID1A* and *APC*, see Methods, Fig. S2, Table S2, Table S3) (14). As expected, somatic tier 1 COSMIC cancer genes were overrepresented (p-value: 4.68×10^-15^, Fig. S3). These results indicate that our methodology can prioritize driver genes purely from functional scores in multiple cancer types.

**Table 1:** Overview of Sequence-based models used in this study.

| Sequence-based model | Reference | Used for | Specifications | Characteristics |
| --- | --- | --- | --- | --- |
| CADDv1.7 (Meta ESM-1v) | (13) | Coding analysis | Pre-computed scores based on a logistic regression model from over 100 annotations, including the Meta ESM-1v Protein language model. | More accurate in coding than in non-coding areas. |
| PARM-K562 & PARM-HepG2 | (8) | Non-coding analysis | Trained on the autonomous promoter potential 600 bp fragments as measured by a reporter assay (Survey of Regulatory Elements) in K562 and HepG2 cell lines. | Smaller model, less computationally intensive. |
| Borzoi | (9) | Non-coding analysis | Trained to simultaneously predict multiple -omics tracks (including RNA-seq) from an input sequence of 524 kbp. | Larger model, more computationally intensive. |

**Table 2:**
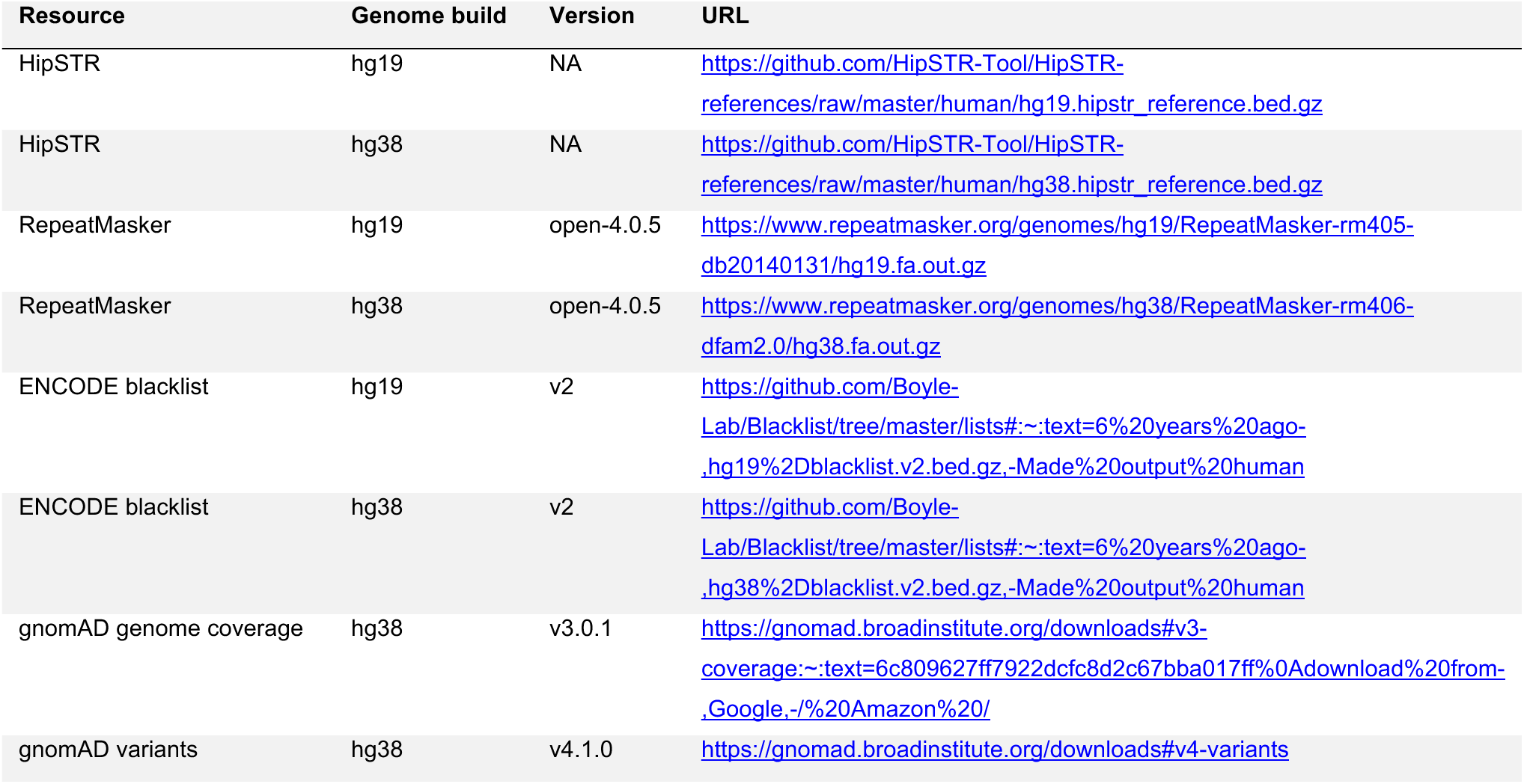
Resources for quality control.

### Functional promoter mutations are significantly enriched in established cancer-driver genes

After confirming our mutational burden methodology with somatic coding SNVs, we applied it to somatic SNVs within non-coding promoter regions of patients in the GEL dataset (see Methods, **Figure 2A**). Briefly, we defined promoter regions as a 1,400 base pair window around a transcription start site (TSS) from refTSS (900 bp upstream and 500 bp downstream) that overlapped an SRTdb transcript observed in either cancer or non-cancer samples (15), resulting in 95,825 regions adjacent to 24,547 genes. We added the 500bp window downstream of the TSS to capture most of the 5’UTR, which may also contain variants that impact expression. We used SRTdb to include all coding and non-coding transcripts, as this database includes transcripts that have been observed in different cancer types but which are not always included in transcript annotation databases such as GENCODE. Similarly, we limited our TSS definition to refTSS to ensure only regions with proven promoter characteristics were included. Variants located within these promoter regions were subjected to additional quality control to remove artifacts, suspected germline variants and variants mapping to coding sequence (CDS) regions. Next, to make inferences about the promoter activity of the remaining somatic SNVs, we separately applied two published PARM sequence-based models (8) trained in two different cell types from a liquid and solid tumour respectively (PARM-K562 and PARM-HepG2, **Table 1**). The predictions from these models are correlated (R=0.54) so we suspect these models can represent other tissues to some extent. PARM is a sequence-based model trained on the autonomous promoter potential of promoter and enhancer 600 bp DNA fragments as measured by a massively parallel reporter assay (promoter-MPRA) (8).

**Figure 2:**
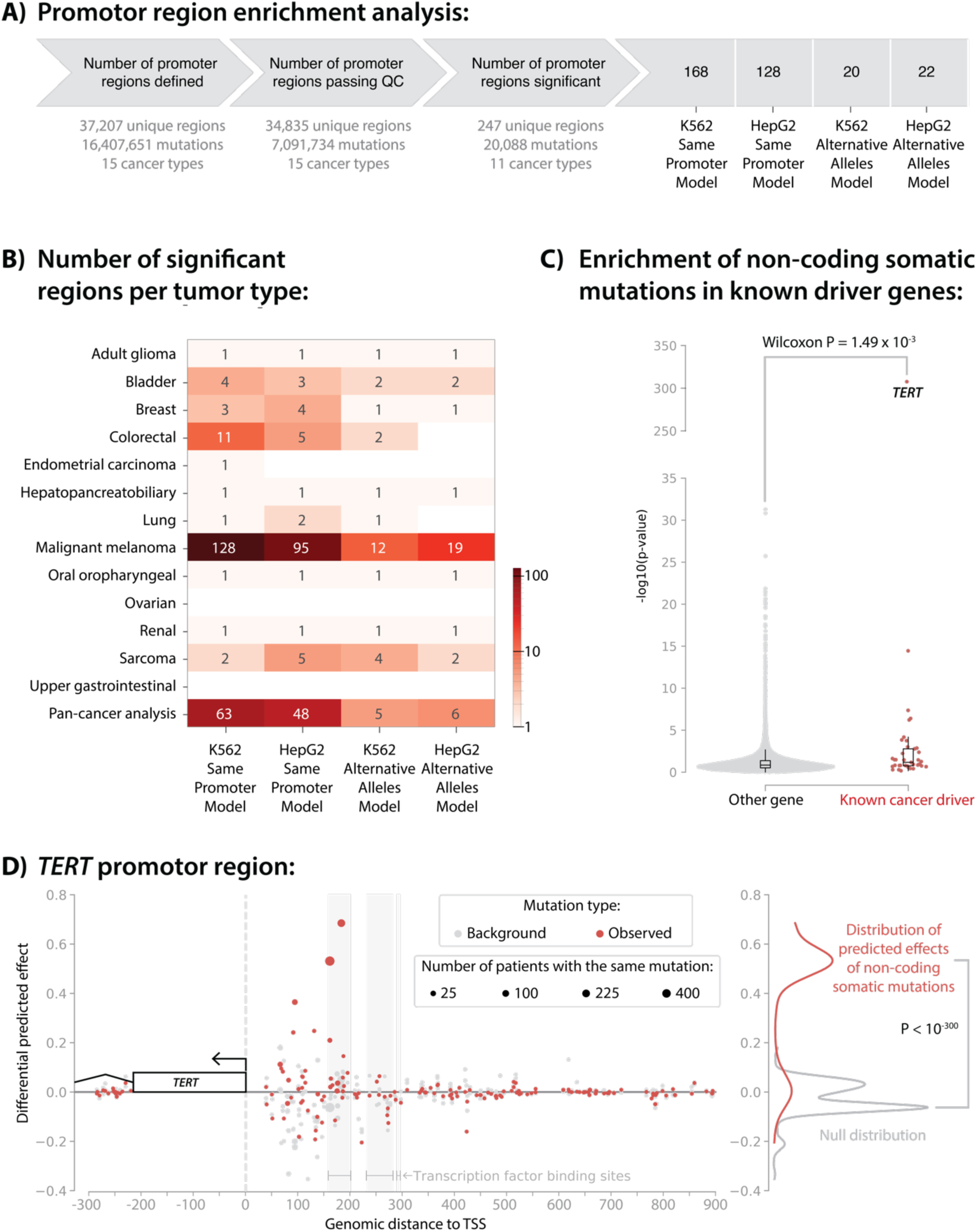
Overview of promoter regions that are enriched for putatively functional non-coding somatic mutations. **A)** Overview of the total number of candidate regions, mutations and cancer types that passed QC and passed Bonferroni-significance (p < 0.05/24,206 regions passing QC = 2.07×10^-6^). **B)** Overview of the mutational burden analysis results: the number of regions (number and colour), cancer types (y-axis) and models (x-axis) considered that were found to be Bonferroni-significant for enrichment of putatively functional non-coding somatic mutations. **C)** Difference of the mutational burden analysis results using PARM between promoter regions of cancer-driver genes and other genes. X-axis shows known cancer-driver genes (in red) and other genes (in light grey). The y-axis shows the -log_10_(p-value) of enrichment of putatively functional non-coding somatic mutations as compared to the background model. **D)** Region plot for the promoter locus of TERT. X-axis indicates distance to the TSS (vertical grey dashed line). Y-axis indicates the differential predicted effect (alternative allele – reference allele) as calculated with the PARM-K562 model. Each red dot indicates an observed somatic non-coding SNV, whereas grey dots indicate predictions for a ‘alternative allele’ background mutation. Dot size indicates the number of patients with that specific somatic variant. Shaded areas show where a saturation in silico mutagenesis experiment using PARM predicts that a transcription motif was disrupted. The distributions of the observed and background mutations are plotted at right. The predicted effects in the observed set are higher than the predicted effects in the background set.

We first applied our mutational burden analysis in a pan-cancer analysis across the entire GEL dataset. After applying quality control and multiple-testing correction, 247 promoter regions passed the Bonferroni threshold when focusing on protein-coding genes (0.05/24,206 regions passing QC and containing >5 variants = 2.07×10^-6^, **Figure 2A**, Table S4), whereas 327 promoters passed Bonferroni when also including non-protein-coding genes (*p* < 0.05/34,835 regions passing QC = 1.44×10^-6^, Table S5). This suggests that enrichment of non-coding variants is not restricted to promotors of coding genes. We also applied our mutational burden analysis per cancer subtype, which showed that most of our significant hits come from melanoma samples (**Figure 2B**; Table S4). To minimize the chance of false positives, we decided to focus our subsequent analyses on promoter regions that could be reproduced across datasets and models. So, for the rest of our analyses, we limited our discovery set to those 247 Bonferroni-significant promoter regions near protein coding genes from the GEL dataset. Of these 247 genes, 18 are known cancer-driver genes (*TERT*, *FGFR2*, *RPS3A*, *CCDC107*, *CDH8*, *ZSCAN5A*, *CSMD3*, *ASXL2*, *TBC1D12*, *TCF3*, *ASPSCR1*, *PRPF40B*, *EBF1*, *TP53*, *PMS2*, *SLC25A46*, *CHEK2* and *PTDSS1*, sorted on significance). The set of 247 genes was also enriched for cancer driver genes overall (Wilcoxon p-value: 1.49×10^-3^, **Figure 2C**), even when excluding *TERT* (Wilcoxon p-value: 3.22×10^-3^). To calculate this enrichment, these results are subset to one result per region, chosen on the lowest p-value over all four combinations of the models (PARM-K562, PARM-HepG2) with background models (alternative alleles, same promoter). Importantly, and reassuringly, this set recapitulates the well-known role of somatic mutations within the promoter region of *TERT*, which had the most significantly prioritized region (p-value < 2.23×10^308^) in our pan-cancer results (**Figure 2D**). Additionally, *TERT* was also individually significant (*p* < 0.05/34,835 regions passing QC = 1.44×10^-6^) in the adult glioma, bladder, hepato-pancreato- biliary, lung, malignant melanoma, oropharyngeal, renal and sarcoma cancer subtype analyses (Table S4). Focusing on subsets of cancer driver types, we observed enrichment for the subset of promotors from tumour suppressor genes (p-value: 6.88×10^-3^), but not for the subset of oncogenes (p-value: 0.09). We suspect this could be because the majority of the SNVs we included in our analysis (74.1%) were predicted by the PARM models to reduce expression, likely favouring the detection of tumour-suppressor promoter regions. This could potentially indicate that non-coding mutations in cancer preferentially regulate cancer driver genes by disrupting transcription factor binding sites.

### 69 promoter regions with enrichment for functional variants are replicated when using independent sequence-based model and datasets

While PARM is trained on MPRA data that measures the autonomous promoter activity of a 600 bp DNA fragment, Borzoi is a large sequence-based model that is trained on epigenetic, chromatin accessibility and gene expression data with a larger window (**Table 1**) (9). Borzoi can make these predictions for multiple different tissues and cell-lines. For all analyses using Borzoi, we took the average prediction over all RNA-seq tissue tracks. Borzoi’s computational intensity (see Methods) makes it less feasible to use this model for analysis of thousands of cancer genomes. We therefore focused on testing whether the 247 unique Bonferroni-significant promoter regions from our PARM-based discovery analysis could be replicated using Borzoi.

Out of the 247 unique Bonferroni-significant regions (identified by any of the PARM-model – background model combinations), 133 (54%) were also significant with Borzoi (*p* < 0.05/ 247 regions of Bonferroni-significance in PARM = 2.02×10^-4^, Fig. S4). For the 247 unique Bonferroni-significant regions, the replication of the direction of effect (predicted increase or decrease in expression), not enforcing replication of significance, was 76.19% using the Same Promoter background model and 66% using the Alternative Alleles background model.

Next, we attempted to replicate the 247 unique Bonferroni-significant regions obtained from the PARM models in GEL using the HMF and ICGC datasets with the PARM models (**Figure 3A**; see Methods). Combined, 156 out of the 247 regions (63%) were significant (*p*<0.05/247=2.0×10^-4^) in HMF after applying either PARM-K562 or PARM-HepG2 using one of either background model (four tests per region). Subsequently, out of these 156 regions, 69 regions were also significant within the ICGC dataset (*p*<0.05/156=3.2×10^-4^) using either PARM model (PARM-K562 or PARM-HepG2) and one of either background model (Same Promoter or Alternative Alleles).

**Figure 3:**
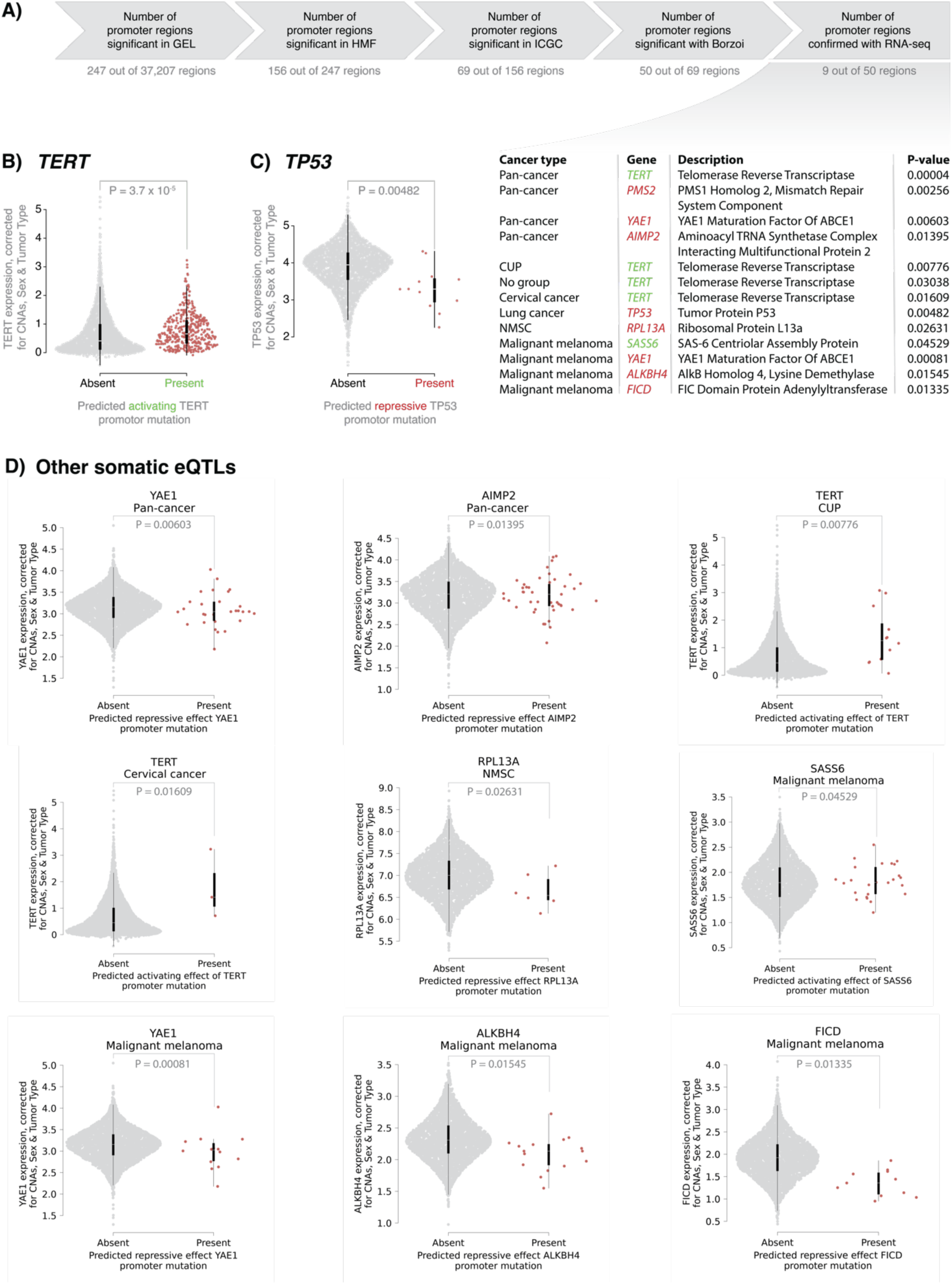
Replication of discovery results in independent datasets, using another sequence-based model and RNA-seq for confirmation. **A)** Upper panel shows an overview of the total number of regions that are Bonferroni-significantly enriched for putative functional non-coding somatic SNVs in GEL (247 regions), that could then be replicated in HMF (156 regions) and the ICGC (69 regions) using both PARM and Borzoi (50 regions) and that could be confirmed with RNA-seq (9 regions). Lower table at right lists all the combinations between cancer type and gene symbols associated with the nine regions that are significantly confirmed using RNA-seq. Gene names are coloured based on their measured activating (green) or repressive (red) effect. CUP = Cancer of Unknown Primary, NMSC = Non-Melanoma Skin Cancer. **B)** Expression of TERT for all cancer patients harbouring mutations predicted by Borzoi to be activating (shown here in red on the right), versus all other cancer patients (shown in grey on the left). Expression has been corrected for copy number alterations, sex, tumour type, tumour purity and expression principal components (PCs) (see Methods) **C)** Expression of TP53 for all cancer patients harbouring mutations predicted by Borzoi to be inactivating/repressive (in red on the right), versus all other cancer patients (shown in grey on the left). **D)** The expression of nine other somatic eQTL – cancer type combination where all cancer patients harbouring mutations predicted by Borzoi to be either repressive or activating (shown here in red on the right), versus all other cancer patients (shown in grey on the left). Expression has been corrected for copy number alterations, sex, tumour type, tumour purity and expression PCs (see Methods).

### High confidence regions that are enriched for functional mutations show matching gene expression changes in paired patient RNA-seq dataset

Next, we attempted to determine whether the predicted effects of our reproducible regions show actual changes in RNA expression levels in cancer patients. For this, we used 3,825 paired RNA-seq samples from the HMF study. To focus on the most reproducible regions, we selected 50 out of the 69 regions that were significant in both the PARM and Borzoi analyses (**Figure 3A**).

We then applied a linear model to fit the RNA-seq expression of the corresponding genes based on the presence or absence of a functional SNV after controlling for sex, tumour purity, copy number state, cancer type and 16 expression principal components. We defined a functional SNV in this context as a SNV of a significant region that is predicted in the same direction of effect, activating or inactivating, that significance is observed in as determined by Borzoi. We subsequently refer to promoter regions that showed altered gene expression in those patients who harbour a predicted functional SNV as “somatic eQTLs” (Methods). Our analysis identified 13 genes with a nominally significant somatic eQTL effect (5x higher than expected by chance, as determined by a binomial test setting the hypothesized probability to 0.05, *p*-value = 9.64 × 10^−7^) and 18 significant somatic eQTL effect – cancer type combinations (2x higher than expected by chance, *p*-value = 3.06 × 10^−6^, **Figure 3A**). Of these 18 somatic eQTL effect – cancer type combinations, 13 cancer-type-specific effects (9 unique genes) showed a direction that was consistent with the PARM and Borzoi predictions that was higher than expected by chance (binomial test *p*-value = 0.048). The low rate of somatic eQTL confirmation (26%; 13/50) may partially be explained by the lack of paired RNA-seq data for all HMF patients studied, with only 3,285 matched RNA-seq samples being available to us.

Reassuringly, we observed a somatic eQTL for the *TERT* promoter, which showed significantly higher (*p*-value: 3.7×10^-5^, beta: 0.13) mRNA expression in patients who harbour promoter SNVs predicted to increase expression when compared to all other cancer patients in HMF (**Figure 3B**). Similarly, we also observed significantly lower expression (*p*-value: 0.00482, beta: -0.44) of the well-known cancer-driver gene *TP53* in patients who harboured SNVs predicted to reduce expression (**Figure 3C**).

Apart from *TERT* and *TP53*, the significant results also included the known cancer driver gene *PMS2*. This gene is a member of the mismatch repair (MMR) pathway often mutated in colorectal cancer (16). In a previous study of tumour genomes with a bigger sample size (*n*=92,439), recurrent non-coding somatic mutations were also identified in the *PMS2* promoter region in skin cancers (17). Within our relatively smaller cohorts (GEL *n*=11,073 samples, HMF *n*=7,171 samples, ICGC n=6,285 samples) we were able to confirm that this region to be enriched for functional variants in melanoma samples (*p*-value: 6.90×10^−7^, **Figure 4A−C**). Since *PMS2* is known to be part of the MMR pathway, this gene has a known functional consequence that can be directly measured in our dataset by determining the mutational burden. Beside the significant repressive eQTL effect (*p*-value: 2.6×10^−3^, beta: -0.14, **Figure 4D**), and in line with the previous report, we also observe a significant higher tumour mutational burden for patients with predicted functional variants in the *PMS2* promotor (*p*-value: 1.0×10^−6^, FC: 3.0, **Figure 4E**). We note however that the effect on mutational burden was much larger than on expression. This suggests that patients with inactivating variants in the promoter of this MMR gene may have a higher total mutational rate over the entire genome. In addition, we also detected a significant somatic eQTL for *AIMP2* (*p*-value: 1.4×10^−2^, beta: -0.12), a gene that is adjacent to *PMS2* on the opposite strand. Since both genes share their promoter region it is possible that this regulatory region may act bidirectionally. *AIMP2* depletion has been shown to induce resistance to apoptosis by interaction with *TP53*, and coding variants that affect this interaction have also been reported (18–20).

**Figure 4:**
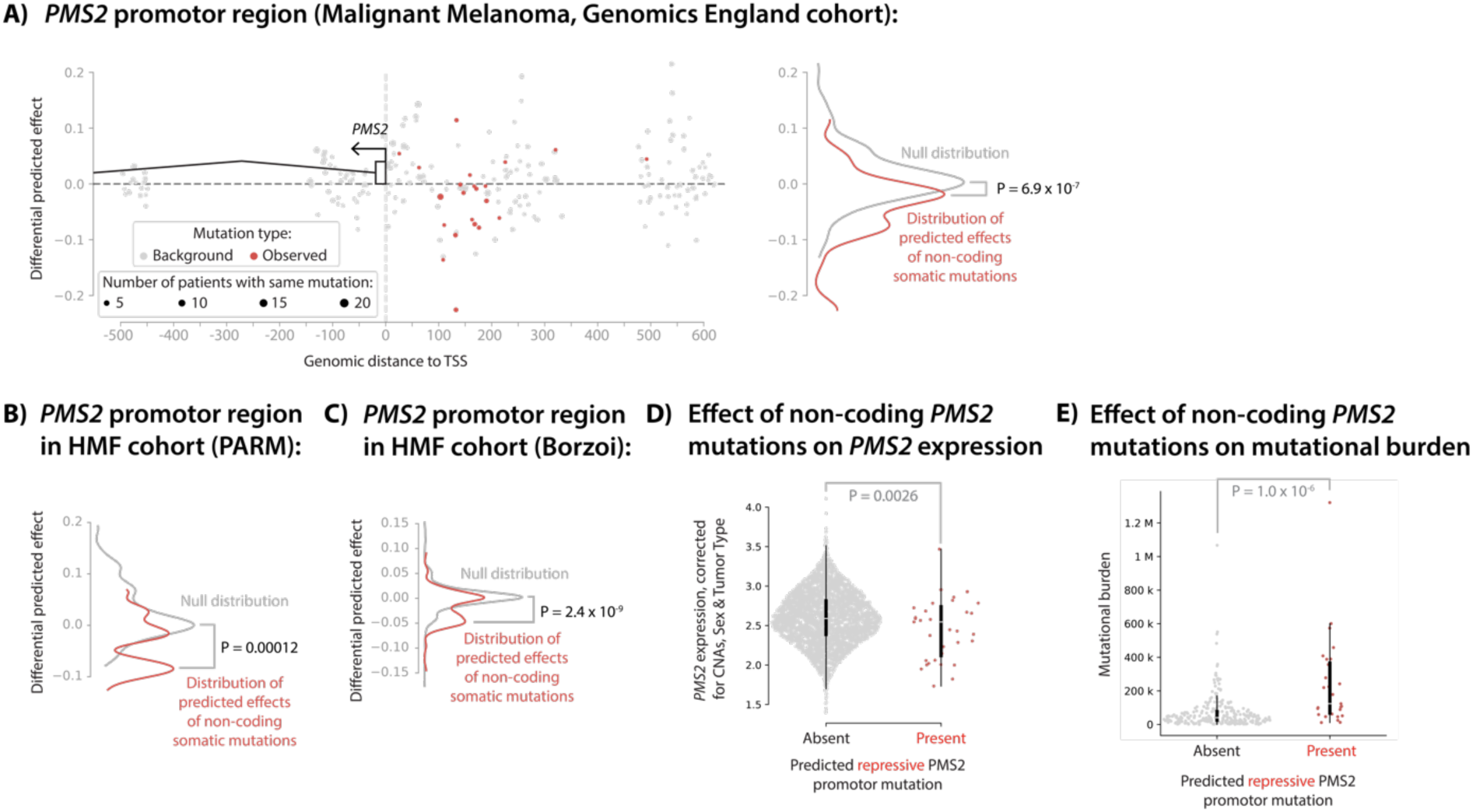
Region plots for the promoter locus of PMS2 and the measured effect of the predicted repressive mutations. **A)** Region plot of the observed and background mutations using PARM-K562 with the Same Promoter background in our discovery set (Genomics England). X-axis shows distance to the TSS (vertical grey dashed line). Y-axis shows the differential predicted effect (alternative allele – reference allele) as calculated with the model (here PARM-K562). Dot size indicates the number of patients with that specific somatic variant. The shaded area’s show where a saturation in silico mutagenesis experiment using PARM predicts a transcription motif is disrupted. The distributions of the observed and background mutations are plotted on the right side. The area-under-the-curve (AUC) of the two-sided Wilcoxon rank-sum test is < 0.5, indicating that the predicted effects in the observed set are lower than the predicted effects in the background set. **B)** Distributions of the observed and background mutations using PARM-K562 with the Same Promoter background in HMF. **C)** Distributions of the observed and background mutations using another independent sequence-based model (Borzoi) with the Same Promoter background in HMF. **D)** Expression of PMS2 of melanoma patients harbouring mutations predicted by Borzoi to be inactivating as shown in panel C (in red on the right), versus all other cancer patients (shown in grey on the left). Expression has been corrected for copy number alterations, sex, tumour type, tumour purity and expression PCs (see Methods) **E)** Number of somatic SNVs (mutational burden) over the entire genome of the same set of melanoma patients as in panel D (red on the right) versus all other melanoma patients (grey on the left).

To our knowledge, this is the first time that functional mutations in non-coding regions have been identified for cancer driver genes other than *TERT*.

Five other candidate promoter regions showed a significant somatic eQTL that had not been reported before, to our knowledge (**Figure 3D**). We observed a significant activating somatic eQTL effect (*p*-value: 4.5×10^−2^, beta: 0.11) for the *SASS6* promotor in melanoma patients. *SASS6* regulates the number of centrosomes in cells during mitosis, and its malfunctioning can result in chromosomal instability (21). *SASS6* overexpression has been associated with oesophageal and lung cancer via *TP53* inhibition (22,23). Reassuringly, we observe an activating somatic eQTL effect that cannot be attributed to amplification since we corrected for copy number with mutations predicted to increase gene expression that also result in higher expression for those patients.

In the non-melanoma skin cancer samples, we observed an inactivating somatic eQTL effect for *RPL13A* (*p*-value: 2.6×10^−2^, beta: -0.33). A same base pair recurrent non-coding somatic mutation at an ETS binding site in the promoter area of *RPL13A* has been observed in melanoma samples before (47 out of 184 melanoma samples) by Frederiksson *et al*. (24).

However, they concluded that this specific mutation does not lead to a change in expression and that the recurrence arises because of a motif preference of UV-induced mutations upstream of CTTCCGG motifs (24). This would suggest that this particular mutation in *RPL13A* is not truly positively selected but rather an artefact of UV damage (24). We also observed this mutation as well in 61 out of 297 melanoma samples, and it is predicted by Borzoi to have only a limited effect on RNA expression (log_2_ differential effect of -0.0021175). However, since we also observed a somatic eQTL at the promotor of *RPL13A* for non-melanoma skin cancer samples, our data suggest there are other functional mutations that can *RPL13A* expression located in the promotor area in non-melanoma skin cancer samples.

We also observed a significant inactivating somatic eQTL effect for *ALKBH4* in melanoma (*p*-value: 1.5×10^−2^, beta: -0.17). *ALKBH4* has been observed to be a tumour promoter in non-small cell lung cancer and a tumour suppressor in colon cancer cell lines (25), the latter being compatible with the inactivating somatic eQTL observed in our study. In colon cancer cell lines, *ALKBH4* overexpression inhibited metastasis through competitive binding of *WDR5* (26). The effect direction of the somatic eQTL we observe is concordant with the promotion of epithelial–mesenchymal transition in our melanoma samples, suggesting that it may play a similar role.

The remaining two genes *FICD* and *YAE1* that showed an inactivating somatic eQTL effect (*p*-value: 1.3×10^−2^ and 8.1×10^−4^, beta: -0.27 and -0.27, respectively) have not been as strongly linked to cancer, making them interesting candidates for future study.

### Meta-analysis in 24,529 tumour WGS samples identifies promoter regions enriched for putatively functional non-coding somatic mutations

In addition to our strict replication approach to identify the most promising candidate driver promoter regions (i.e. sequentially enforcing Bonferroni significance in GEL, ICGC and HMF), we also performed a pan-cancer meta-analysis (on the WGS data of all 24,529 samples) using both PARM-K562 and PARM-HepG2 (see Methods). This identified a set of 492 Bonferroni- significant genes Table S1) that is significantly enriched for known cancer driver genes (p- value: 9.70×10^-5^, **Figure 5A**). These genes also show significantly higher evolutionary constraint (higher intolerance to loss-of-function variation), which we observed through a lower 90% CI upper bound of the observed vs. expected score from gnomAD (27) (one-sided Wilcoxon rank-sum test *p*-value: 2.76×10^-5^, FC: 0.88, **Figure 5B**). We then used ToppGene (28) to ascertain the enrichment of predicted gene functions and observed Bonferroni-significant enrichment for the Gene Ontology Biological Process terms ‘mitotic cell cycle’, ‘mitotic cell cycle process’, ‘cell cycle’ and ‘cell cycle process’ were significantly enriched (Bonferroni-adjusted p-values of 7.67×10^-5^, 1.13×10^-4^, 1.50×10^-3^ and 2.21×10^-3^ respectively, **Figure 5C**). Furthermore, the set of 492 genes is also strongly enriched for certain transcription factor targets such as genes targeted by ELF2, DIDO1 and SETD1A (Bonferroni-adjusted p-values of 8.50×10^-61^, 3.65×10^-34^, and 2.50×10^-24^ respectively). This may suggest either preferential transcription factor dysregulation in cancer or, potentially, a technical bias of the sequence-based models we have applied here.

**Figure 5:**
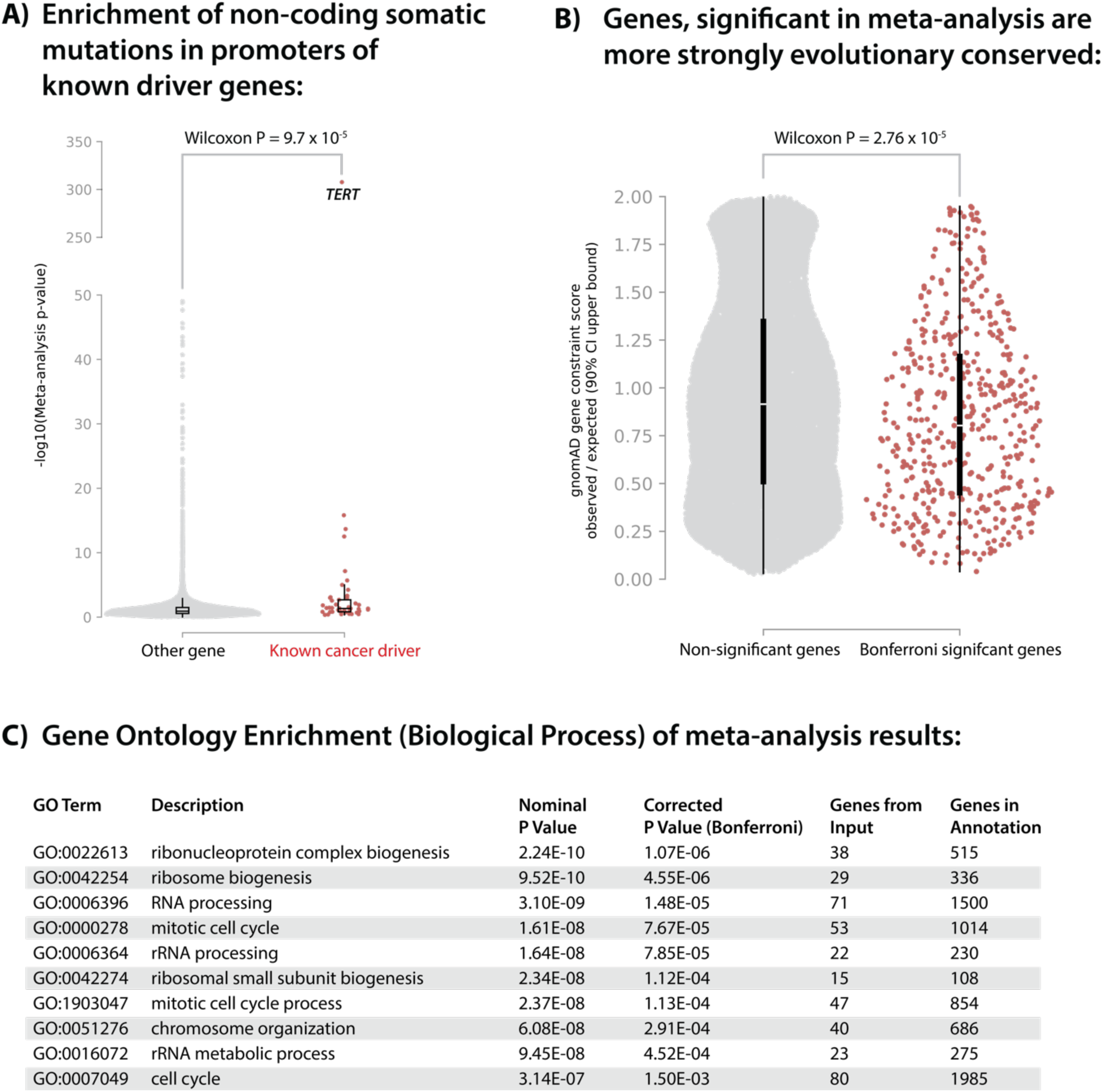
Meta-analysis enrichments. **A)** Significance of the mutational burden analysis results using PARM on promoter regions of cancer-driver genes and other genes within the meta-analysis between GEL, HMF and ICGC. X-axis shows the known cancer-driver genes (red) and other genes (light grey). Y-axis shows the -log_10_(meta-analysis p-value) of enrichment of putatively functional non-coding somatic mutations as compared to the background model. These results are subset to one result per region, chosen based on the lowest p-value over all four combinations of the models (PARM-K562, PARM-HepG2) with background models (Alternative Alleles, Same Promoter). Cancer-driver genes showed significantly lower p-values than other genes (p-value: 9.7·10^5^). **B)** Evolutionary constraint as measured by gnomAD’s observed / expected (oe) score. X-axis shows the Bonferroni significant genes from the meta-analysis (in red) and other genes (in light grey). Y-axis shows the lower 90% CI upper bound of the observed/expected score. The lower observed/expected score(p-value: 2.76·10^5^) for Bonferroni significant genes from the meta-analysis indicates significantly more genomic constraint. **C)** Bonferroni significant GO: biological process terms identified through ToppGene.

Overall, we consider that this set of 492 Bonferroni-significant genes from the meta-analysis more accurately approximates the number of promoter regions that show evidence of putatively functional enrichment with our methodology than an analysis in a single dataset. We believe these findings could be valuable as a resource for the scientific community (Table S1).

## Discussion

In this study, we have shown that sequence-based model predictions can be leveraged to prioritize non-coding regions enriched for putatively functional variants in cancer. We show an enrichment of known cancer-driver genes near these prioritized regions. After applying a stringent combination of filters (replication in three independent cohorts, confirmation by Borzoi and overlap and concordance with somatic expression QTL) we identified nine candidate driver promoter regions, that includes those for well-known cancer-driver genes such as *TERT*, *TP53* and *PMS2*. However, the results we obtained with less stringent filter combinations suggest that many more promoter mutations could be cancer drivers, and meta-analysis of the three combined cohorts identified 483 additional regions that are enriched for putatively functional non-coding somatic mutations that could also be of interest. We observed that genes in these regions are more evolutionary constrained, are enriched for being involved in the cell-cycle and are more often established cancer driver genes. Follow-up research is warranted to clarify the role of these genes in carcinogenesis.

We can make these inferences by first using sequence-based models to predict the direction of effect of individual somatic mutations and by subsequently testing whether an excess of either activating or repressive effects are observed in a region of interest. Earlier work, such as that from the PCAWG Consortium could not yet take advantage of these direction predictions.

We believe this is one of the reasons why these earlier studies concluded that non-coding somatic driver mutations were rare, whereas we find evidence this is likely a quite common phenomenon. Our observations are consistent with recent genome-wide association studies in cancer that have uncovered hundreds of genetic risk factors in non-coding regions of the genome (2).

Within the enriched regions we observe that non-coding somatic mutations preferentially have an expression-decreasing effect, likely because a random mutation within a transcription factor binding site will more easily lead to lower binding affinity than that it will increase it. In line with this we also observe an enrichment of established tumour suppressor genes within such regions. We observed that the somatic eQTLs have small effect sizes, which is in line with what has been observed for germline eQTLs before as well (29–31). One explanation for this is that eQTL effects can be highly tissue and cell-type specific and that strong effects only manifest in a subset of the cells that comprise the studied tumour biopsies (29–31). Therefore, we reason that the effect sizes we described for non-coding functional mutations could be underestimated.

However, this does not mean that these variants could not have large downstream effects. This is illustrated by the *PMS2* example: even though the observed somatic eQTL was small (beta: - 0.14), we observed a much stronger effect on the tumour mutational burden (FC: 3.0). Follow-up work should determine whether specific mutational signatures that are associated to DNA mismatch repair deficiency are also *increasing* in patients harbouring these putatively inactivating mutations in the promoter of *PMS2*.

Our analysis identified many known tumour suppressor genes, but not all of them. As such, we believe our sample size is still not near saturation. With the planned inclusion of more tumour WGS in the future the power to detect these variants will increase. Integrating more paired RNA-seq profiles will also be valuable: out of the 24,529 studied WGS tumour samples, we had only 3,825 matched RNA-seq tumour samples available to us. This would maximize statistical power to validate the expression changes of non-coding variants in current WGS profiles.

We believe future analyses will also benefit from faster, tissue or cell type-specific models that can also make inferences on distant regulatory regions. We have restricted our non-coding analyses to the promoters, which only cover a limited amount of the non-coding genome. To discover further relevant non-coding cancer mutations, future work should include the analysis of distal regulatory elements, with the challenge lying in interpreting the target gene and the uncertainty of interpreting the effect size.

In conclusion, we have shown that non-coding somatic mutations can be prioritized for functional effect using sequence-based models, leading to the identification of a substantial number of candidate cancer promotor regions that could be of future interest for diagnostics and drug targeting. This work showcases a methodology to study the non-coding tumour genome, especially when sample size is limited or for hypermutated tumour types. We hope that with the improvement of sequence-based models, strategies like the methodology we propose here will help further unravel the functional landscape of non-coding variation within the tumour genome.

## Methods

### Selection of non-coding regions centred around TSS

We anchored the promoter regions of this study around TSSs from the refTSS v4.1 data set (32). To focus on TSSs relevant to cancer cells, we overlapped the TSS peaks with the 5’ end of RNA-seq-derived cancer transcripts defined by the SRTdb (accessed on 7^th^ of February 2024) (15). We included all transcripts defined in the SRTdb (CCLE, GTEx and TCGA). We observed that some transcripts missed their refTSS peak by a few basepairs. To account for inaccurate transcript boundaries in SRTdb we extended the 5’ end of SRTdb transcripts in increments of 1 bp and determined the increase in additional overlapping regions. We chose a 5’ extension of 22 bps after we observed that for more than 22 bps, the increase in additional captured regions was smaller than the increase when adding the first base pair. After determining a cancer-relevant refTSS subset, we created a 900-bp upstream and 500-bp downstream window around the middle of the peak to define our promoter regions. We fixed the end of a region as 500 bp downstream of the middle of the refTSS peak because this is the 98% percentile of the length distribution of the 5’ untranslated regions (5’UTRs) of protein-coding genes. This resulted in 95,825 regions adjacent to 24,547 genes.

### QC to limit false somatic non-coding mutation calls

The GEL (hg38) and HMF (hg19) datasets use a different pipeline for somatic variant-calling that includes filters from variants found in matched normal tissue. For GEL and HMF we included variants that were annotated with a ‘PASS’ in the QC column. For ICGC we included all single base pair substitution variants derived from WGS. Despite the extensive QC of the somatic variant-calling pipelines of GEL, HMF and ICGC, we still observed there were recurrent SNVs annotated with high frequency (filtering allele frequency; FAF > 0.01% gnomAD v4.1) in some ancestries (27). In addition, some SNVs were located within short tandem repeats, low complexity regions and homopolymer repeats. It is difficult to determine if a variant is a SNV change or a insertion/deletion which are difficult to predict with current sequence-based models. Alternatively, a mutation that was called as somatic could also be a sequencing artefact. To prevent our mutational burden methodology from prioritizing regions based on suspected germline variants and possible sequencing artefacts, we applied seven additional filters:

1. To avoid artefacts from regions that are difficult to sequence we removed all SNVs located within genomic regions defined by the ENCODE blacklist (version 2) (33).
2. To remove putative germline variants, we removed all SNVs reported as a variant that passed QC in Exomes or Genomes of gnomAD v4.1.0 when they had a FAF (95% CI GroupMax FAF) of > 0.01%.
3. We removed SNVs located within a base pair position that was reported with low coverage in gnomAD (genome coverage summary TSV v3.0.1 with < 30x coverage).
4. We removed SNVs that were located within a simple repeat or low complexity region as reported by RepeatMasker (RepeatMasker open-4.0.5 – Repeat Library 20140131) (34).
5. We removed SNVs located within a homopolymer repeat of at least 5 bps.
6. We removed SNVs located within an single tandem repeat STR mapped by HipSTR for repeats runs of at least five repetitions of a repeat unit of 1-5 bps (35).
7. Finally, we removed all SNVs within a coding region (CDS) according to Gencode (v45). When studying exon regions with CADD1.7, this step was skipped.

Only regions with more than five mutations were considered for the mutational burden analysis.

For all resources that did not have a hg19 equivalent we used Python 3.9.1 with the liftover 1.2.2 package to create hg19 equivalent files from the hg38 resources . This resulted in 34,835 regions that pass QC.

### Mutational burden testing

Since single somatic SNVs generally have a low recurrence across individuals, we developed a burden analysis framework that evaluates all observed SNVs in a promoter region at once. To calculate a specific promoter burden, we first determine a list of all observed SNVs passing QC across all individuals in the discovery dataset. Then, for each SNV, we predict the differential effect on gene expression using a sequence-based model (i.e.: CADD for coding regions and PARM-K562, PARM-HepG2 or Borzoi otherwise). Additionally, for each prediction, we devise a matching null prediction generated using a background model (i.e.: the “Alternative alleles” model or the “Same Promoter” model). Finally, we test the difference between the observed and null predictions as follows: we create a distribution of predicted values across the real observed SNVs and a distribution for the matched null predictions and test the difference between these distributions using a Wilcoxon signed rank test, yielding a p-value for the tested region. To be able to reasonably perform our enrichment analysis between observed mutations and mutations in our null distribution we enforce that a region should harbour mutations from at least 5 different donors. We performed these analyses for different discovery datasets within the Genomics England dataset: over all cancers (pan-cancer) and per cancer subtype.

### Strategies for creation of mutational background tests

To test if the distribution of predictions of the putative functional effect of non-coding SNVs is higher or lower than expected by chance, we set out to create two different mutational background tests (Fig. S1).

Firstly, to prioritize regions that contain SNVs that are predicted to increase expression via specific gain-of-function variants (such as the *TERT* promoter), we designed the Alternative Alleles background test. In this test, we compare the observed mutations against the non-observed alternative bp changes at the same base pair position. For example, if the observed allele change at a given position is A>T, we compare this against all other possible allele changes at that position (A>G and A>C) as a null background. When a background variant is also observed in the same or another patient within that cohort, we exclude it from our background set. This test then prioritizes variants where the observed allele change has an absolute predicted effect that is larger than the other two potential allele changes. This could potentially be the case when a motif gets created, such as the ETS motif in the *TERT* example.

The second background set was designed to prioritize regions that contain SNVs that are predicted to have an effect regardless of allele change because of their sequence context within the promoter. For example, this could be the case when a transcription motif gets disrupted. We created this null background model by again determining the predictions for base pair changes, but this time at positions elsewhere in the same promoter that share the same trinucleotide context of the variant before the somatic mutation (hence the name Same Promoter). As with the Alternative Alleles background test, when the background variant was observed in the same or another patient within that cohort, we exclude it from our background set.

### Prediction of the consequence of coding somatic SNVs using CADD

To show that our methodology and background tests can prioritize cancer-drivers purely from prediction scores without modelling the background mutational rate of the tumour, we first applied our mutational burden analysis to coding variants (retaining the CDS regions during QC). We parsed the GEL dataset for coding mutations falling within Gencode v45 canonical transcript exons between 56 and 1403 bp in length (36). We scored these coding SNVs with pre-computed predictions (“raw scores”) from CADD1.7 derived from the sequence-based model Meta ESM-1v (13). To boost statistical power, we jointly tested all exon regions of the same gene rather than of each exon individually.

### Prediction of the differential gene expression effect of non-coding somatic SNVs with PARM and Borzoi

Because it would would take at least 2,738 hours (or 114 days) of compute time to run Borzoi for all defined regions, we decided to first run our mutational burden analysis with the PARM model genome-wide across the 34,835 regions that passed QC (8,9). In comparison to Borzoi’s speed of 0.71 variants per second, PARM can score 435 variants per second (measured on an NVIDIA A40 GPU). For PARM, we used the K562- and HepG2-based models. The models were developed as described in PARM (8), but with a filter size of 300 instead of 125 and a slight modification in the fragment selection process for training. In addition to using fragments overlapping promoters, as described in the original PARM paper, we also included fragments overlapping enhancer regions, as defined by ChromHMM (37). Unless stated otherwise, for the analyses we describe here we subset the data to regions linked to protein-coding genes only from Gencode v41, resulting in 24,206 regions passing QC associated with protein-coding transcripts. We apply multiple hypothesis correction calculated by dividing the alpha of 0.05 by the number of regions considered in an analysis to limit false positives. We subsequently replicated only on the regions that our pipeline calls Bonferroni-significant (threshold = 2.07×10^-6^, *p*-value < 0.05/24,206 regions passing QC), and tested for significant replication with Borzoi (*p* < 0.05/247 regions of Bonferroni-significance in PARM = 2.02×10^-4^). For the Borzoi predictions score, we used the mean over all the GTEx RNA tracks (over all the available tissues) for the gene annotated to our promoter region.

### Collection of cancer-driver genes from previous studies

To compare our results to known cancer-driver genes we collected previously identified drivers from the COSMIC Cancer Gene Census (version 96, obtained from the official website in May 2022) (38), the IntOGen catalogue (release 2020.02.01) (1) and Dietlein *et al*. (4), and we have made these available in the supplements (Table S3). The Dietlein *et al.* results were obtained from the supplementary files provided with their publication. We call a driver gene “cancer-specific” when it was called in association with that cancer type in any of the three collections. To harmonize the gene identifiers, we processed all the gene symbols and identifiers from these collections into Ensembl gene identifiers using the “MAPIDs” function in the R package “org.Hs.eg.db”. We label a gene as an oncogene or a tumour suppressor based on COSMIC’s “oncogene” and “TSG” labels or for IntOGen the “Act” and “LoF” labels.

### Enrichment analyses in results of mutational burden analysis

We tested if the *p*-values of enrichment for putatively functional non-coding somatic mutations are associated with cancer-specific driver status (matched on tissue-of-origin and cancer type). Since a region could be significant as result of a specific combination of background models, pan-cancer or cancer type sub-selection and PARM model, we included all results within our discovery set (GEL) using any of the combination between PARM and background model and selected the observation with the lowest *p*-value for each individual region. We then compared the cancer gene’s minimal *p*-values vs other genes’ minimal *p*-values using one-sided Wilcoxon rank-sum test. To show that this enrichment is not driven by the *TERT* locus, we repeated this analysis after while removing promoter regions overlapping *TERT* transcripts, before applying the one-sided Wilcoxon rank-sum test again.

### Replication analysis in the HMF and the ICGC datasets

To replicate our findings in other independent datasets we applied our mutational burden analysis to the HMF and the ICGC datasets. Pan-cancer comparisons between GEL, HMF and ICGC datasets were made including all samples derived from WGS from non-metastatic tissue without balancing the subpopulations of cancer types in both datasets. To compare cancer-specific types across cohorts we manually created a conversion table between the cancer types defined in GEL, HMF and the ICGC (Table S6).

### Processing of RNA-seq data in HMF

To confirm the predictions of non-coding somatic mutations on gene expression, we processed RNA-seq data matched to WGS samples. For this, we used the HMF dataset (*n*=7,171 WGS samples) since this dataset includes tumour-matched RNA-seq for 3,825 samples. We took the adjusted TPM counts from Isofox at HMF (https://github.com/hartwigmedical/hmftools/tree/master/isofox) and removed all samples with an adjusted TPM count > 2^23^ to discard outliers. Furthermore, we kept only samples that had non-zero expression values for > 40% of the tested genes. Finally, we removed genes that had zero expression in > 20% of the samples. We then log_2_-transformed, centred and scaled the data such that the mean = 0 and the standard deviation = 1. (Fig. S5).

### Somatic eQTL analysis

Inspired by the work of Pudjihartono *et al.*, we set out to confirm our replicating findings that are enriched for putatively functional non-coding somatic SNVs using RNA-seq (39). This “Somatic eQTL” is modelled as a multivariate regression model using ordinary least squares that models expression based on the mutational status of a promoter region of a patient in combination with multiple covariates as follows:

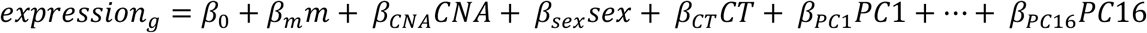

Where *expression_g_* is the log_2_(adjTPM) of gene *g*, *β*_0_ represents the intercept and *β_m_m* is the mutational status, defined as 0 if the patient has no mutation in the direction of effect (enrichment in activating or inactivating predictions) of AUC of the two-sided Wilcoxon rank-sum test and as 1 if the patient has at least one mutation in the direction of effect of AUC of the two-sided Wilcoxon rank-sum test. *β_CNA_CNA* is a constant for the median copy number alteration, taken from the minimum and maximum copy number alteration for that gene defined by HMF. *β_sex_sex* is set to 0 for female, and 1 for male. *β_CT_CT* is a one-hot encoded for the 27 cancer types defined in HMF. *β*_*PC*1_*PC*1 through *β*_*PC*1_*PC*1 are the expression principal components (PCs). The 16 PCs where chosen based on the 16 hidden factors used by Pudjihartono *et al.* (39).

### Tumour mutational burden analysis for *PMS2*

In addition to our confirmation of putatively functional mutations on expression, we also looked at confirmation on gene function level. We chose to focus on *PMS2* to study the functional consequences of the putatively functional mutations because we could compare the tumour mutational burden since *PMS2* plays a role in DNA mismatch repair (16,17). For this, we took all the non-coding somatic mutations from Genomics England located in the promoter region of *PMS2* that were predicted by Borzoi to have a negative differential effect. Next, we took the pre- computed number of SNVs as shared within the Genomics England Research Environment by Kinnersley *et al.* (40), and compared these between melanoma patients who harboured at least one of the mutations predicted to be inactivating and all other melanoma patients who do not harbour these mutations.

### Meta-analysis of the GEL, HMF and the ICGC datasets

To boost statistical power and identify other potential candidate driver promoter regions we used a meta-analysis strategy to combine pan-cancer and cancer-type-specific results between GEL, HMF and ICGC. We manually determined which cancer type is present in multiple datasets and created a conversion table (Table S6). We transformed the p-values to Z-scores and calculated a combined Z-score using the weighted Z-method as described by Whitlock (41), where we use the square root over the number of samples in a cancer subtype as the weights.

## Declarations

### Ethical Compliance

This research project has been reviewed by the UMCG medical ethical committee under project number 20440 and has been deemed as non-WMO research.

All cohorts included in this study enrolled participants with informed consent and collected and analyzed data in accordance with ethical and institutional regulations. Information about individual institutional review board approvals is available in the original publications for each cohort. Where applicable, data access agreements were signed by the investigators prior to acquisition of the data to the UMCG which state the data usage terms. To protect the privacy of the participants, data access was restricted to the investigators of this study, as defined in those data access agreements. Per data use agreements, only summary level data is made publicly available and strictly mentioned in the disclaimer that they cannot be used to re-identify study participants.

### Data availability

Our study is comprised of previously published cancer datasets. These datasets are available upon request, or through online repositories after signing data access agreements. We have listed the mode of access for each of the included datasets below.

#### Genomics England

Genomics England 100,000 Genomes Project data was requested through the Genomics England Research Portal under the research registry ID “1125” (https://research.genomicsengland.co.uk/).

#### Hartwig Medical Foundation

Hartwig Medical Foundation data was requested through under the unique ID “DR-413”.

#### International Cancer Genome Consortium

International Cancer Genome Consortium data has initially been accessed through the ICGC data portal (https://dcc.icgc.org/) which officially closed in June 2024, after which we requested access again at ICGC ARGO (https://www.icgc-argo.org) under the DACO application # “DACO-2065”.

### Code availability

Code is available at: https://github.com/curzuat/nocosomu_iDriver/tree/dev_tvl.

### Author contributions

- Conceptualisation: T.v.L., C.G.U.T., L.F.
- Data curation: T.v.L., C.G.U.T.
- Formal Analysis T.v.L., C.G.U.T.
- Funding acquisition: L.F., B.v.S., J.d.R., E.V.
- Investigation: T.v.L., C.G.U.T., M.C.L.B ,L.F.
- Methodology: T.v.L., C.G.U.T., L.B.M., N.H.M.K., V.H.F.S., H.J.W., J.d.R., L.F.
- Software: T.v.L., C.G.U.T., L.B.M.
- Supervision: C.G.U.T, L.F.
- Visualisation: T.v.L., L.F.
- Writing – original draft: T.v.L., C.G.U.T., L.F.
- Writing – review & editing: T.v.L., C.G.U.T., L.B.M., N.H.M.K., V.H.F.S., M.C.L.B, M.P., H.J.W., B.v.S., J.d.R., E.V., L.F.

Roles as defined by: CRediT (Contributor Roles Taxonomy)

## Acknowledgments

This publication and the underlying study have been made possible partly based on data that Hartwig Medical Foundation has made available to the study through the Hartwig Medical Database.

The data for this study are provided (in part) by the Netherlands Cancer Institute (Antoni van Leeuwenhoek ziekenhuis). The authors want to thank all the involved patients, medical staff and research staff for making this possible.

This research was made possible through access to data in the National Genomic Research Library, which is managed by Genomics England Limited (a wholly owned company of the Department of Health and Social Care). The National Genomic Research Library holds data provided by patients and collected by the NHS as part of their care and data collected as part of their participation in research. The National Genomic Research Library is funded by the National Institute for Health Research and NHS England. The Wellcome Trust, Cancer Research UK and the Medical Research Council have also funded research infrastructure.

We also would like thank the ICGC data access committee for providing access to variant calls used in this study, the patients that agreed to participate as part of the ICGC project, the clinical networks that coordinated the submission of data to ICGC, and the data access committee for providing access to variant calls used in this study.

Research at the Netherlands Cancer Institute is supported by an institutional grant of the Dutch Cancer Society and of the Dutch Ministry of Health, Welfare and Sport. The Oncode Institute is partially funded by the Dutch Cancer Society.

We would like to thank Kate Mc Intyre and Monique van der Wijst for editorial assistance and feedback on this manuscript. We would like to thank the Center for Information Technology of the University of Groningen for their support and for providing access to the Habrok high-performance computing cluster, as well as the UMCG Genomics Coordination center, the UG Center for Information Technology, and their sponsors BBMRI-NL and TarGet for storage and compute infrastructure.

L.F. is supported by a grant from the NWO (ZonMW-VICI 09150182010019 to L.F.), through a Senior Investigator Grant from the Oncode Institute, a grant from Oncode Accelerator and a grant from Saxum Volutum (Pericode). Aditionally, B.v.S., J.d.R. and E.V. are also supported through a Senior Investigator Grant from the Oncode Institute and a grant from Saxum Volutum (Pericode).

## Supplementary data

**Fig. S1.**
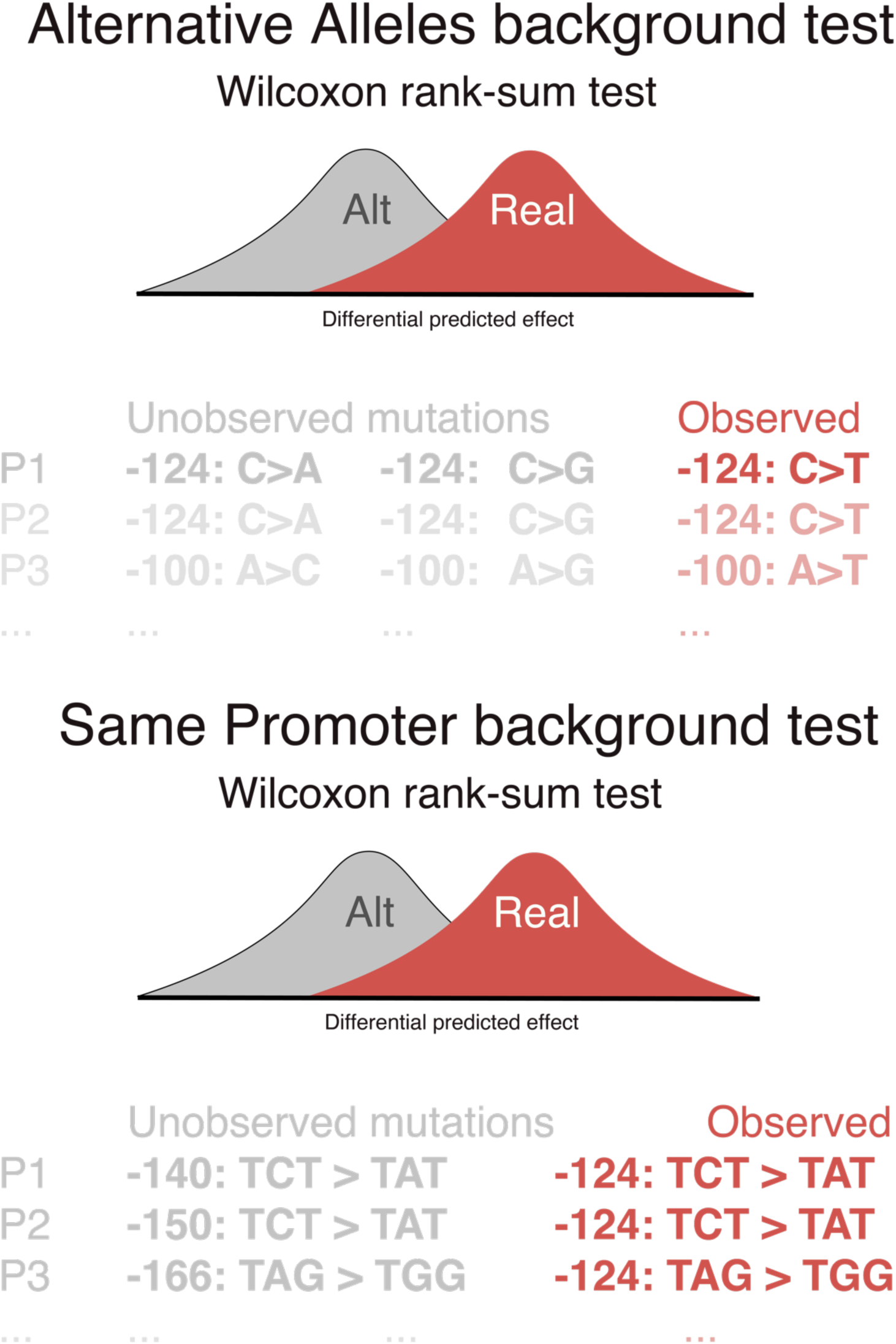
Illustration showing the creation of the Alternative Alleles and Same Promoter background mutation tests.

**Fig. S2.**
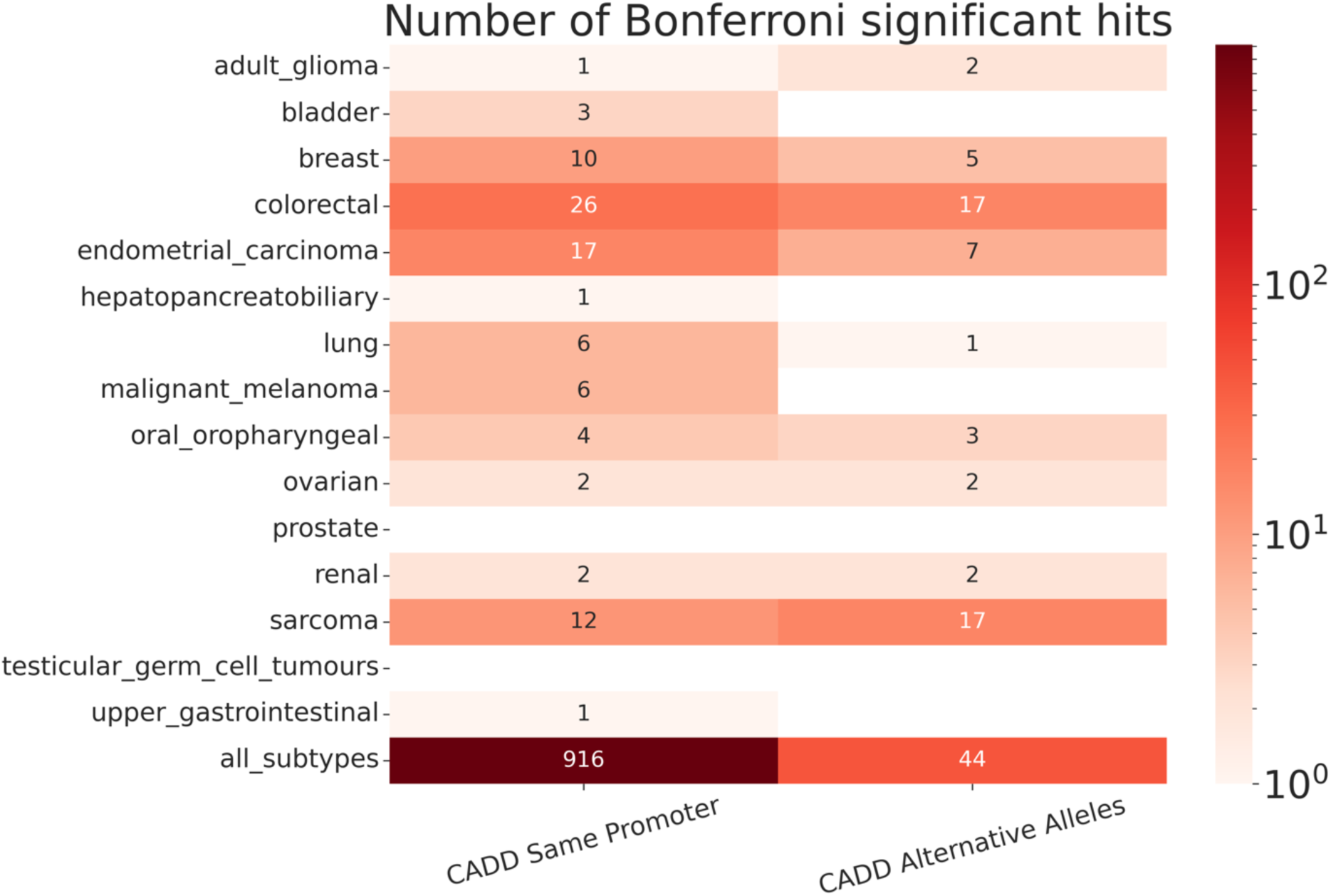
Overview of the mutational burden analysis results using CADD on the number of exons (number and colour), cancer types (y-axis) and models (x-axis) considered that were found to be Bonferroni-significant for enrichment of putatively functional coding somatic mutations.

**Fig. S3.**
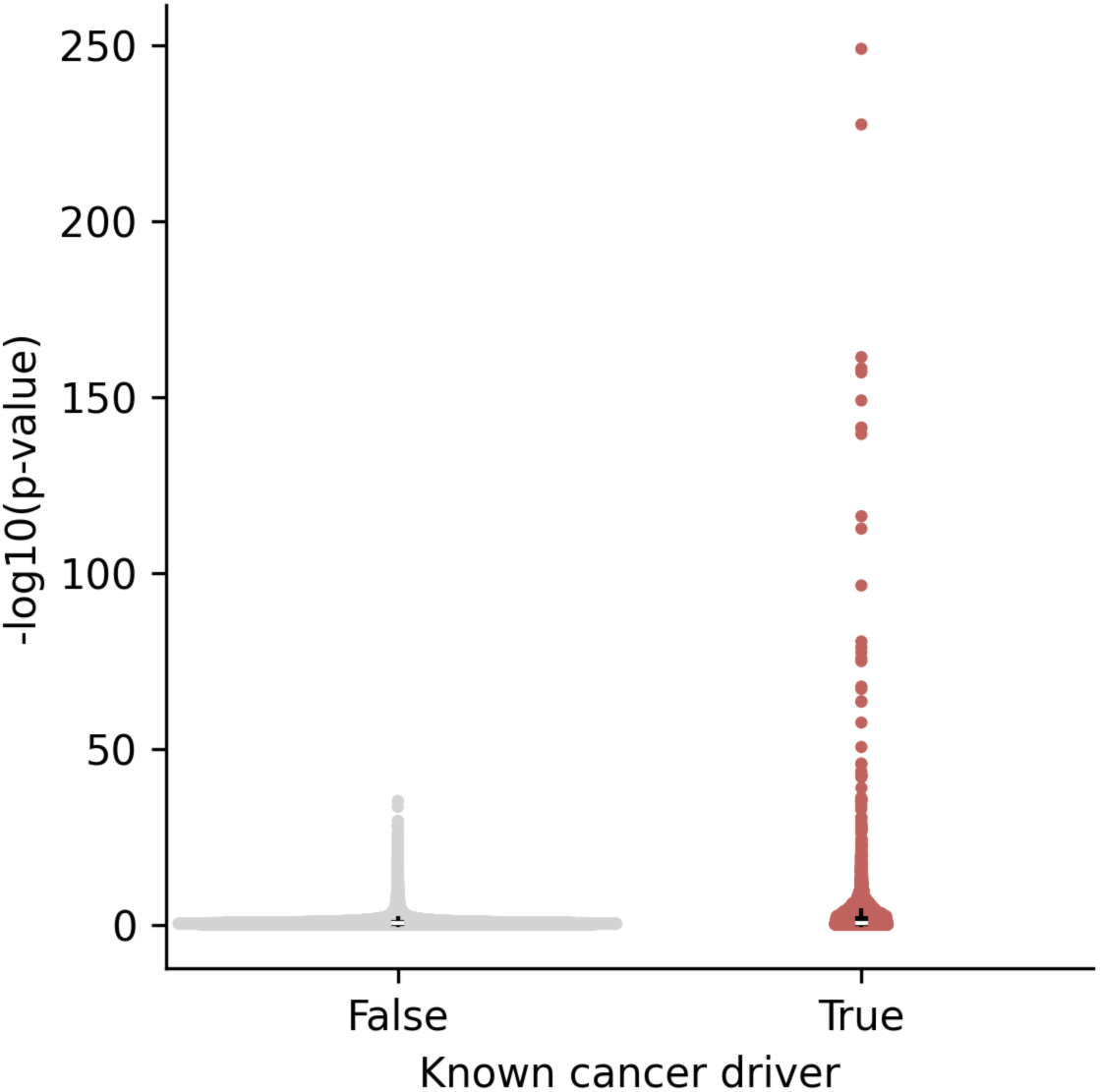
Significance of known cancer drivers vs. other genes for the coding region analysis using CADD.

**Fig. S4.**
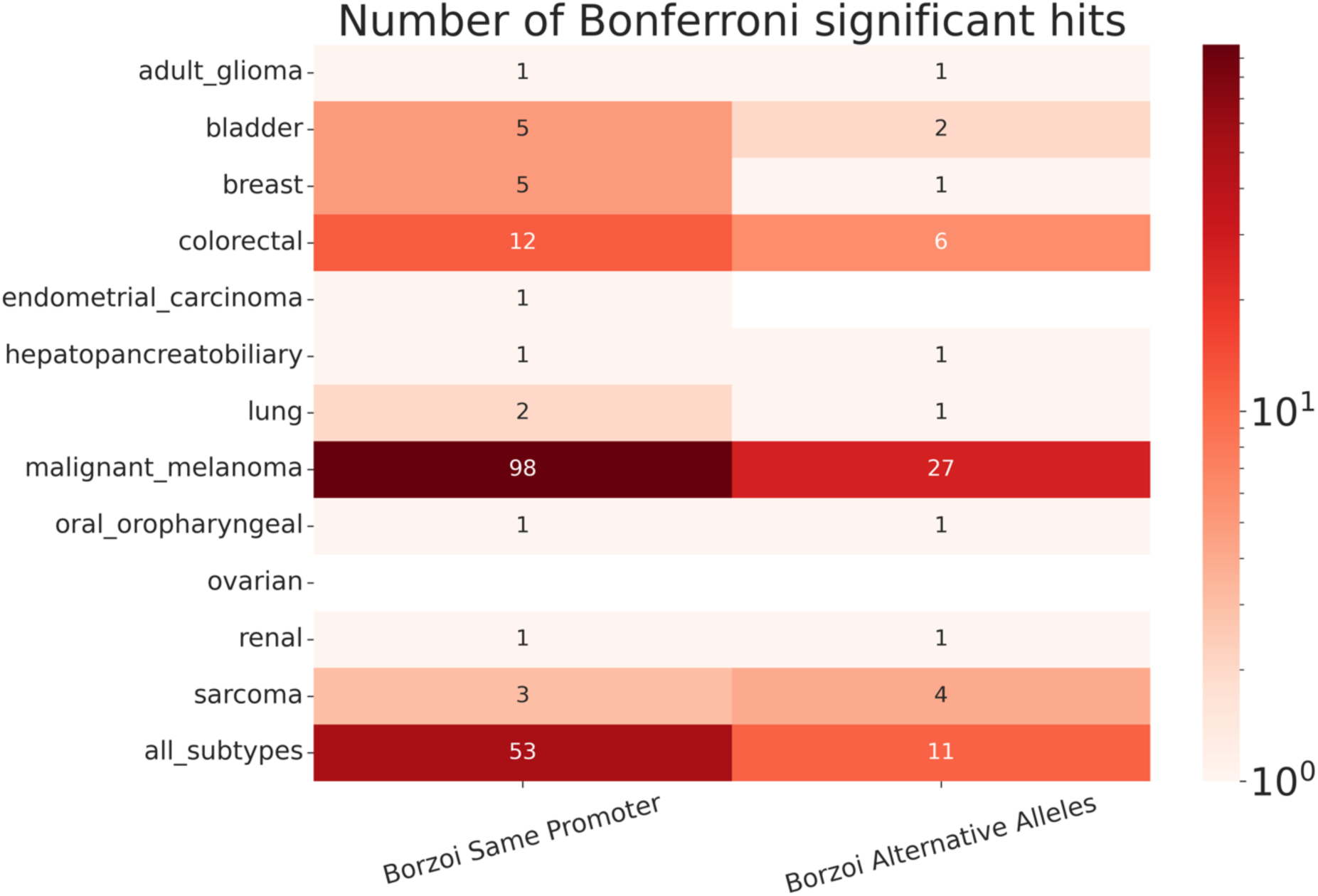
Overview of the mutational burden analysis results using Borzoi on the number of regions (number and colour), cancer types (y-axis) and models (x-axis) considered that were found to be Bonferroni-significant for enrichment of putatively functional coding somatic mutations.

**Fig. S5.**
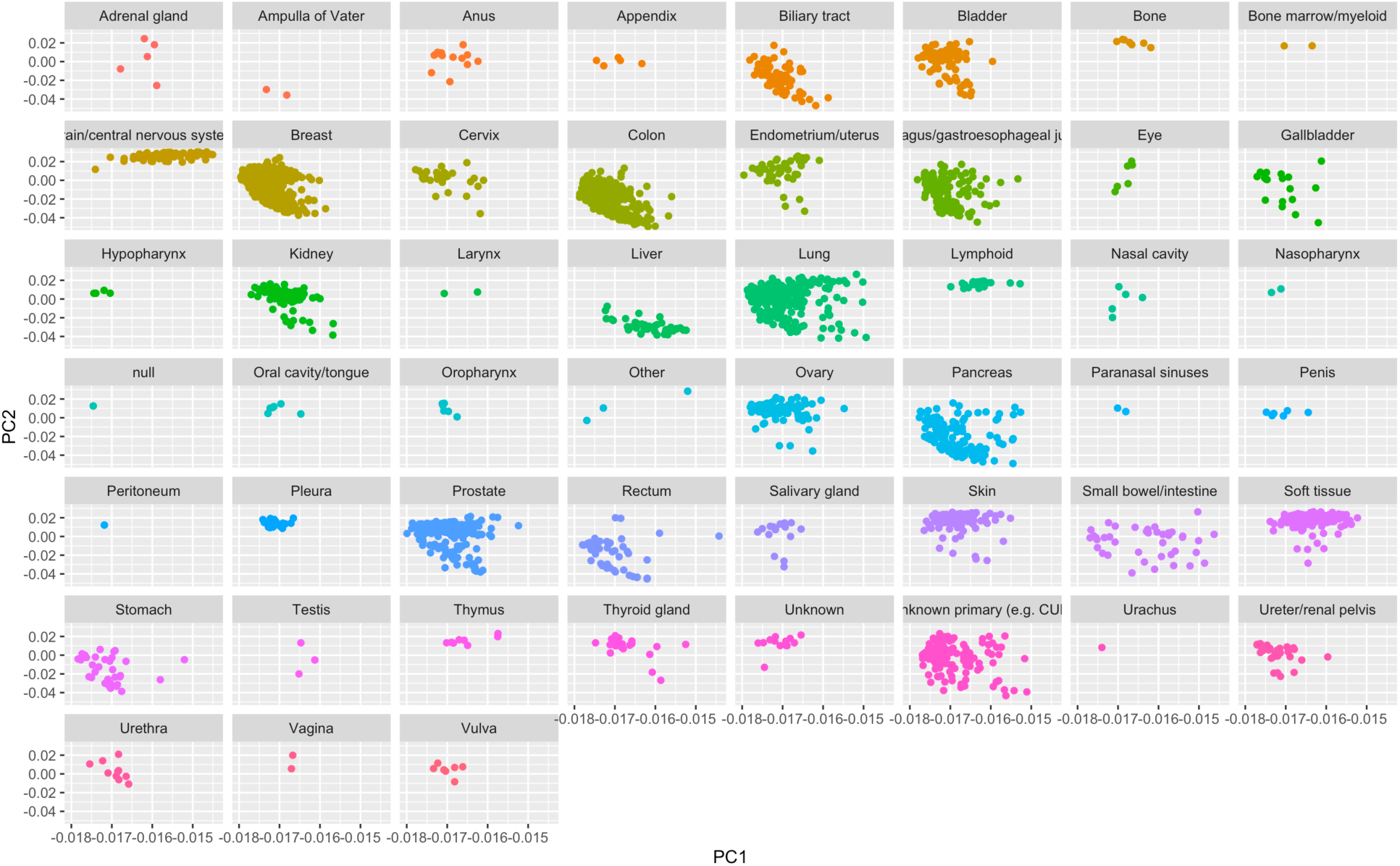
Visual inspection of expression principal components of Hartwig Medical Foundation RNA-seq samples.

Table S1. Meta-analysis results on promoters of protein-coding genes of Genomics England, Hartwig Medical Foundation and the International Cancer Genome Consortium

Table S2. Mutational burden analysis results on protein-coding genes using CADD on aggregated transcript level annotated by known cancer-driver status

TableS3. Cancer-driver gene collection

Table S4. Mutational burden analysis results on promoter of protein-coding genes using PARM annotated by known cancer-driver status

Table S5. Mutational burden analysis results on promoters, including promoters of protein- coding genes and non-coding RNAs using PARM annotated by known cancer- driver status

Table S6. Conversion table of cancer types between Genomics England, Hartwig Medical Foundation and the International Cancer Genome Consortium

## References

1. Martínez-Jiménez F, Muiños F, Sentís I, Deu-Pons J, Reyes-Salazar I, Arnedo-Pac C, et al. A compendium of mutational cancer driver genes. Nat Rev Cancer. 2020 Oct;20(10):555– 72.

2. Fanfani V, Citi L, Harris AL, Pezzella F, Stracquadanio G. The Landscape of the Heritable Cancer Genome. Cancer Res. 2021 May 15;81(10):2588–99.

3. Rheinbay E, Nielsen MM, Abascal F, Wala JA, Shapira O, Tiao G, et al. Analyses of non- coding somatic drivers in 2,658 cancer whole genomes. Nature. 2020;578(7793):102–11.

4. Dietlein F, Wang AB, Fagre C, Tang A, Besselink NJM, Cuppen E, et al. Genome-wide analysis of somatic noncoding mutation patterns in cancer. Science. 2022 Apr 8;376(6589):eabg5601.

5. Avsec Ž, Agarwal V, Visentin D, Ledsam JR, Grabska-Barwinska A, Taylor KR, et al. Effective gene expression prediction from sequence by integrating long-range interactions. Nat Methods. 2021 Oct;18(10):1196–203.

6. Karollus A, Mauermeier T, Gagneur J. Current sequence-based models capture gene expression determinants in promoters but mostly ignore distal enhancers. Genome Biol. 2023 Mar 27;24(1):56.

7. Dudnyk K, Cai D, Shi C, Xu J, Zhou J. Sequence basis of transcription initiation in the human genome. Science. 2024 Apr 26;384(6694):eadj0116.

8. Barbadilla-Martínez L, Klaassen N, Franceschini-Santos VH, Breda J, Hernandez-Quiles M, Lieshout T van, et al. The regulatory grammar of human promoters uncovered by MPRA- trained deep learning [Internet]. bioRxiv; 2024 [cited 2024 Dec 15]. p. 2024.07.09.602649. Available from: https://www.biorxiv.org/content/10.1101/2024.07.09.602649v2

9. Linder J, Srivastava D, Yuan H, Agarwal V, Kelley DR. Predicting RNA-seq coverage from DNA sequence as a unifying model of gene regulation. Nat Genet. 2025 Jan 8;1–13.

10. The National Genomic Research Library v5.1, Genomics England. 10.6084/m9.figshare.4530893.v7.

11. Stichting Hartwig Medical Foundation. Hartwig Medical Foundation. [cited 2025 Feb 7]. Database with genetic and clinical information of cancer patients. Available from: https://www.hartwigmedicalfoundation.nl/en/

12. Zhang J, Bajari R, Andric D, Gerthoffert F, Lepsa A, Nahal-Bose H, et al. The International Cancer Genome Consortium Data Portal. Nat Biotechnol. 2019 Apr;37(4):367–9.

13. Schubach M, Maass T, Nazaretyan L, Röner S, Kircher M. CADD v1.7: using protein language models, regulatory CNNs and other nucleotide-level scores to improve genome- wide variant predictions. Nucleic Acids Res. 2024 Jan 5;52(D1):D1143–54.

14. Urzúa-Traslaviña CG, van Lieshout T, Boulogne F, Domanegg K, Zidan M, Bakker OB, et al. Co-expression in tissue-specific gene networks links genes in cancer-susceptibility loci to known somatic driver genes. BMC Med Genomics. 2024 Jul 15;17(1):186.

15. Shi Q, Liu T, Hu W, Chen Z, He X, Li S. SRTdb: an omnibus for human tissue and cancer- specific RNA transcripts. Biomark Res. 2022 Apr 26;10(1):27.

16. ten Broeke SW, van der Klift HM, Tops CMJ, Aretz S, Bernstein I, Buchanan DD, et al. Cancer Risks for PMS2-Associated Lynch Syndrome. J Clin Oncol. 2018 Oct 10;36(29):2961–8.

17. Chalmers ZR, Connelly CF, Fabrizio D, Gay L, Ali SM, Ennis R, et al. Analysis of 100,000 human cancer genomes reveals the landscape of tumor mutational burden. Genome Med. 2017 Apr 19;9(1):34.

18. Kim DG, Lee JY, Lee JH, Cho HY, Kang BS, Jang SY, et al. Oncogenic Mutation of AIMP2/p38 Inhibits Its Tumor-Suppressive Interaction with Smurf2. Cancer Res. 2016 May 31;76(11):3422–36.

19. Han JM, Park BJ, Park SG, Oh YS, Choi SJ, Lee SW, et al. AIMP2/p38, the scaffold for the multi-tRNA synthetase complex, responds to genotoxic stresses via p53. Proc Natl Acad Sci. 2008;105(32):11206–11.

20. Choi JW, Kim DG, Park MC, Um JY, Han JM, Park SG, et al. AIMP2 promotes TNFα- dependent apoptosis via ubiquitin-mediated degradation of TRAF2. J Cell Sci. 2009;122(15):2710–5.

21. Leidel S, Delattre M, Cerutti L, Baumer K, Gönczy P. SAS-6 defines a protein family required for centrosome duplication in C. elegans and in human cells. Nat Cell Biol. 2005 Feb;7(2):115–25.

22. Li Z, He L, Li J, Qian J, Wu Z, Zhu Y, et al. SASS6 promotes tumor proliferation and is associated with TP53 and immune infiltration in lung adenocarcinoma. Clin Exp Med. 2024 Oct 24;24(1):243.

23. Xu Y, Zhu K, Chen J, Lin L, Huang Z, Zhang J, et al. SASS6 promotes proliferation of esophageal squamous carcinoma cells by inhibiting the p53 signaling pathway. Carcinogenesis. 2021 Feb 1;42(2):254–62.

24. Fredriksson NJ, Elliott K, Filges S, Eynden JV den, Ståhlberg A, Larsson E. Recurrent promoter mutations in melanoma are defined by an extended context-specific mutational signature. PLOS Genet. 2017 May 10;13(5):e1006773.

25. Shen C, Yan T, Tong T, Shi D, Ren L, Zhang Y, et al. ALKBH4 Functions as a Suppressor of Colorectal Cancer Metastasis via Competitively Binding to WDR5. Front Cell Dev Biol [Internet]. 2020 May 14 [cited 2025 Feb 13];8. Available from: https://www.frontiersin.org/journals/cell-and-developmental-biology/articles/10.3389/fcell.2020.00293/full

26. Li MM, Nilsen A, Shi Y, Fusser M, Ding YH, Fu Y, et al. ALKBH4-dependent demethylation of actin regulates actomyosin dynamics. Nat Commun. 2013 May 14;4(1):1832.

27. Chen S, Francioli LC, Goodrich JK, Collins RL, Kanai M, Wang Q, et al. A genomic mutational constraint map using variation in 76,156 human genomes. Nature. 2024 Jan 1;625(7993):92–100.

28. Chen J, Bardes EE, Aronow BJ, Jegga AG. ToppGene Suite for gene list enrichment analysis and candidate gene prioritization. Nucleic Acids Res. 2009 Jul 1;37(suppl_2):W305–11.

29. THE GTEX CONSORTIUM. The GTEx Consortium atlas of genetic regulatory effects across human tissues. Science. 2020 Sep 11;369(6509):1318–30.

30. de Klein N, Tsai EA, Vochteloo M, Baird D, Huang Y, Chen CY, et al. Brain expression quantitative trait locus and network analyses reveal downstream effects and putative drivers for brain-related diseases. Nat Genet. 2023 Mar;55(3):377–88.

31. Võsa U, Claringbould A, Westra HJ, Bonder MJ, Deelen P, Zeng B, et al. Large-scale cis- and trans-eQTL analyses identify thousands of genetic loci and polygenic scores that regulate blood gene expression. Nat Genet. 2021 Sep;53(9):1300–10.

32. Abugessaisa I, Noguchi S, Hasegawa A, Kondo A, Kawaji H, Carninci P, et al. refTSS: A Reference Data Set for Human and Mouse Transcription Start Sites. J Mol Biol. 2019 Jun 14;431(13):2407–22.

33. Amemiya HM, Kundaje A, Boyle AP. The ENCODE Blacklist: Identification of Problematic Regions of the Genome. Sci Rep. 2019 Jun 27;9(1):9354.

34. Smit, AFA, Hubley, R. & Green, P. RepeatMasker [Internet]. Available from: http://www.repeatmasker.org

35. Willems T, Zielinski D, Yuan J, Gordon A, Gymrek M, Erlich Y. Genome-wide profiling of heritable and de novo STR variations. Nat Methods. 2017 Jun;14(6):590–2.

36. Mudge JM, Carbonell-Sala S, Diekhans M, Martinez JG, Hunt T, Jungreis I, et al. GENCODE 2025: reference gene annotation for human and mouse. Nucleic Acids Res. 2025;53(D1):D966–75.

37. Ernst J, Kellis M. Chromatin-state discovery and genome annotation with ChromHMM. Nat Protoc. 2017 Dec;12(12):2478–92.

38. Sondka Z, Bamford S, Cole CG, Ward SA, Dunham I, Forbes SA. The COSMIC Cancer Gene Census: describing genetic dysfunction across all human cancers. Nat Rev Cancer. 2018 Nov;18(11):696–705.

39. Pudjihartono M, Pudjihartono N, O’Sullivan JM, Schierding W. Melanoma-specific mutation hotspots in distal, non-coding, promoter-interacting regions implicate novel candidate driver genes. Br J Cancer. 2024 Nov;131(10):1644–55.

40. Kinnersley B, Sud A, Everall A, Cornish AJ, Chubb D, Culliford R, et al. Analysis of 10,478 cancer genomes identifies candidate driver genes and opportunities for precision oncology. Nat Genet. 2024 Sep;56(9):1868–77.

41. Whitlock MC. Combining probability from independent tests: the weighted Z-method is superior to Fisher’s approach. J Evol Biol. 2005 Sep 1;18(5):1368–73.

